# Sex differences in the influence of toxoplasmosis on stress and perceived anxiety: Evidence for the stress-coping hypothesis

**DOI:** 10.1101/2022.01.24.22269770

**Authors:** Jaroslav Flegr, Šárka Kaňková

**Affiliations:** Division of Biology, Faculty of Science, Charles University, Prague, Viničná 7, Prague 128 44, Czech Republic

**Keywords:** *Toxoplasma gondii*, toxoplasmosis, anxiety, stress, stress-coping hypothesis

## Abstract

Life-long infection with *Toxoplasma*, which affects 30% of the human population, has specific behavioral effects. The stress-coping hypothesis explains why the toxoplasmosis-associated behavioral changes go in opposite directions in men and women. It suggests that toxoplasmosis impairs the health of humans, which results in chronic stress. Men and women are known to cope with stress in opposite ways. The first presumption of the hypothesis, impaired health, was confirmed in many studies. The second, higher level of stress, was tested only rarely. Levels of stress and anxiety, measured with the Perceived Stress Scale, and the State-Trait Anxiety Inventory X-2, respectively, were compared in a population of 614 *Toxoplasma*-free and 162 *Toxoplasma*-infected subjects. Higher stress was detected in the infected men, but not women. We also found that physical health had a positive rather than negative effect on stress when mental health is controlled, which seems to contradict the prediction of the stress-coping hypothesis. No differences were found in the anxiety of infected and noninfected subjects. Subjects who have objective reasons for stress (those with worse physical health) are less stressed than those without such reasons.

## 1. Introduction

Latent toxoplasmosis, the lifelong infection by a protozoan parasite *Toxoplasma gondii*, affects about one-third of the world population (Tenter, Heckeroth, & Weiss, 2000). It is associated with the occurrence of specific personality and behavioral traits, such as low novelty-seeking conscientiousness, higher emotional warmth in women, and lower warmth and higher suspiciousness in men, for review see, e.g., (Lindová, Příplatová, & Flegr, 2012). The intensity of many of these changes increases with time passed since the infection (Flegr, Kodym, & Tolarová, 2000; Flegr, Zitkova, Kodym, & Frynta, 1996), suggesting that they represent the effects of the latent infection, not fading effects of a more dramatic but short phase of acute disease.

To explain the existence of many cumulative personality changes and especially to explain why these changes mostly go in the opposite direction in men and women, Lindová *et al*. suggested the so-called stress-coping hypothesis (Lindová, et al., 2010; Lindová, et al., 2006). According to this hypothesis, the latent *Toxoplasma* infection is associated with impaired health in infected subjects, which results in mild but long-term chronic stress. It is known that a sex-specific behavioral response to stress exists in humans. While men employ more individualistic and antisocial (e.g., aggressive, hostile) forms of coping (Carver, Scheier, & Weintraub, 1989; Hobfoll, Dunahoo, Ben-Porath, & Monnier, 1994), women under stress are more likely to seek and provide social support (Carver, et al., 1989; Rosario, Shinn, Mørch, & Huckabee, 1988; Stone & Neale, 1984), join with others (Hobfoll, et al., 1994), and verbalize towards others or the self (Tamres, Janicki, & Helgeson, 2002). All behavioral changes that go in the opposite direction in the *Toxoplasma*-infected men and women, e.g., decreased extroversion, cooperativeness, and strength of superego in men and increase of these traits in women (Flegr & Hrdý, 1994; Flegr, et al., 1996) can be explained in this theoretical framework.

The first presumption of the stress-coping hypothesis, namely impaired health of the subjects with latent toxoplasmosis, has been already confirmed in many studies (reviewed in (Flegr, Prandota, Sovickova, & Israili, 2014)). However, the second presumption, the increased level of stress in *Toxoplasma*-infected subjects, has been tested only rarely. The only data suggesting a higher level of stress measured with a psychological instrument (DASS21 questionnaire) showed that the *Toxoplasma*-infected subjects scored higher in stress and probably also anxiety (Shirbazou, Abasian, & Meymand, 2011). The infected subjects also expressed an increased level of the stress hormone cortisol and testosterone. This cross-sectional study was, however, performed on a relatively small population of 180 subjects, all of them being non-specified patients of Sina Hospital in Tehran. Another study showing an increased level of cortisol in *Toxoplasma*-infected subjects was performed in Sudan (Abdelazeem, Moddawe, Ahmed, & Abdrabo, 2015). In this study, one hundred infected subjects, patients of a military hospital in Khartoum, had about two times higher concentrations of cortisol than 50 *Toxoplasma*-free controls (634 vs 324 nmol/l). In contrast, no significant difference in the concentration of cortisol (but a trend in the right direction p = 0.077) between infected and non-infected patients was observed in 120 schizophrenia patients in Egypt (El-Gebaly, et al., 2019).

A typical emotional response to long, mild, unpredictable stress is anxiety (Endler & Parker, 1990). Many studies have shown the association between toxoplasmosis and the level of anxiety or the incidence of generalized anxiety disorder (Akaltun, Kara, & Kara, 2018; Alvarado-Esquivel, et al., 2016; Bak, et al., 2018; Flegr & Horáček, 2020; Groer, et al., 2011; Markovitz, et al., 2015; Suvisaari, Torniainen-Holm, Lindgren, Harkanen, & Yolken, 2017). However, even here some contradictory results exist (Gale, Brown, Berrett, Erickson, & Hedges, 2014; Markkula, Lindgren, Yolken, & Suvisaari, 2020).

The main purpose of the present study was to search for further evidence in favor or against the stress-coping hypothesis, namely evidence of the increased level of perceived stress in subjects infected with *Toxoplasma*. We used an electronic survey containing Perceived Stress Scale, and the State-Trait Anxiety Inventory X-2 to collect personality and anamnestic data from a large sample of the nonclinical internet population individuals that were laboratory tested for toxoplasmosis.

## 2. Material and methods

### 2.1 Subjects

The participants of the present study were recruited from members of the Facebook and internet community Labbunnies and their Facebook friends using the Facebook-based snowball method (Kankova, Flegr, & Calda, 2015). They were invited to do an internet test to learn how anxious and stressed they are in comparison to other people. At the end of the questionnaire, they were informed that their data can be also used for scientific purposes – the study of the relation between certain biological factors, namely toxoplasmosis, contact with pets, and natural hair color with anxiety and stress, and were provided the opportunity to ask for erasing their data. Less than 1% asked for this. All subjects got the information about their results in the tests and the subjects with a very high level of anxiety or stress (top 5% quantile) were recommended to seek professional help. The study was conducted following relevant guidelines and regulations.

### 2.2 Questionnaire

The questionnaire was operated on the Qualtrics platform. In the anamnestic part of it, the respondents answered the questions about their age, sex, toxoplasmosis status, and Rh phenotype (it is known for a long time that Rh-positive subjects are much more resistant to adverse effects of latent toxoplasmosis than Rh-negative subjects, for review see (Flegr, Novotná, Lindová, & Havlíček, 2008; Flegr, Toman, Hula, & Kankova, 2020)). Namely, they were asked to respond whether they had been laboratory tested for toxoplasmosis and the result of this test (negative/positive-infected/I do not know, I am not sure). They were reminded that toxoplasmosis is a parasite of cats that is especially dangerous for pregnant women. The third response “I do not know, I am not sure” was checked in advance. Similarly, they were asked about their Rh blood group (positive/negative/I do not know, I am not sure). They were reminded that Rh-negative blood is the rarer variant, and the third response was checked in advance. The participants were also asked to rate their physical conditions and mental conditions using two six-point scales anchored with 1 – bad, 6 – excellent. The main parts of the questionnaire were the Czech version of the 20-item self-report scale for anxiety symptoms State-Trait Anxiety Inventory X-2 (STAI X-2) (Heretik, Ritomský, Novotný, Heretik, & Pečeňák, 2009), and 10-item self-report Perceived Stress Scale (PSS) as a classic stress assessment instrument (Cohen & Williamson, 1988). STAI X-2 measured anxiety as a trait, respondents read the statements and choose how they feel generally. Answers are recorded on a 4-point scale (almost never, sometimes, often, almost always). The overall score may range from 20 to 80, with a higher score indicating greater anxiety. PSS finds out how often the respondent perceived the surrounding events in the last month as unpredictable, uncontrollable, and stressful. Answers are recorded on a 5-point scale (never, almost never, sometimes, quite often, very often). Each item can contribute 0 to 4 points to an overall score, so the resulting score ranges from 0 to 40, with a higher score indicating a higher intensity of perceived stress.

### 2.3 Statistics

Between 28. 9. 2020 and 13. 4. 2021, the questionnaire was completed or partially completed by 6000 subjects, 5200 of them during October and November 2020. We filtered out all subjects who were younger than 18, did not provide information about toxoplasmosis or Rh phenotype, and skipped more than four questions in STAI or more than two questions in PSS, or answered nearly all questions in STAI or PSS with the same code. We calculated average anxiety and stress as the arithmetic means of answers of STAI and PSS questionnaires. These means were multiplied by the number of questions in a particular questionnaire (20 in the case of STAI and 10 in the case of PSS) to allow comparison to published data. The final data set contained data from 776 subjects who mostly completed both questionnaires. The set was uploaded to the public repository Figshare doi.org/10.6084/m9.figshare.14651490.v1 (Flegr, 2021).

The effect of toxoplasmosis on anxiety and perceived stress was assessed both with parametric (MANCOVA and ANCOVA) and nonparametric methods. Primarily (for making our conclusions), we used the nonparametric method partial Kendall correlation, which is not sensitive to the distribution of output variables, differences in the number of subjects in particular groups, and the presence of outliers. Partial Kendall correlation was computed in R 3.3.1 (R Core Team, 2018) using the package ppcor (Kim, 2015) and our package of scripts Explorer 1.0 (https://doi.org/10.6084/m9.figshare.14685825.v1) (Flegr & Flegr, 2021). All other tests and descriptive statistics were done with the statistical package Statistica v. 10.0.

## 3. Results

### 3.1 Descriptive statistics

The final sample consisted of 776 subjects, 614 *Toxoplasma*-free (mean age 34.18, S.D. 11.37) and 162 *Toxoplasma*-infected (mean age 39.29, S.D. 10.57), the difference in age was significant (*t*_774_ = -5.16, *p* < 0.0001). The difference in age between 138 *Toxoplasma*-free (mean age 34.96, S.D. 12.90) and 18 *Toxoplasma*-infected men (mean age 40.17, S.D. 9.34) was not significant (*t*_154_ = -1.66, *p* = 0.100), while the difference in age between 476 *Toxoplasma*-free (mean age 33.95, S.D. 10.89) and 144 *Toxoplasma*-infected women (mean age 39.01, S.D. 10.75) was highly significant (*t*_627_ = -5.06, *p* < 0.0001). The prevalence of toxoplasmosis was 11.54% in men and 23.23% in women; this difference was again highly significant (*Chi*^2^ = 10.31, *p* = 0.0013). In contrast, the difference in the frequency of Rh-negativity between men 23.08% and women 22.10% was not significant (*Chi*^2^ = 0.07, *p* = 0.793). The prevalence of toxoplasmosis was approximately the same in 173 Rh-negative (20.81%) and 603 Rh-positive subjects (20.90%, (*Ch*i^2^ < 0.01, *p* = 0.980).

### 3.2 Influence of toxoplasmosis on anxiety and stress

The association between toxoplasmosis and the dependent variables anxiety and stress was studied using multivariate analysis of covariance (MANCOVA) with toxoplasmosis, sex, Rh, and interactions toxoplasmosis-sex, Rh-toxoplasmosis, and sex-Rh as independent factors and age as a covariate. Only the effects of age (*F*_2,761_ = 26.04, *p* < 0.0001) and toxoplasmosis-sex (*F*_2,761_ = 3.84, *p* = 0.022) were significant. Very similar results were provided in simplified models without toxoplasmosis-sex, or toxoplasmosis-sex and Rh-sex interactions, or even the simplest model with only sex, toxoplasmosis, and toxoplasmosis-sex interaction as the independent factors and age as a covariate (age: *F*_2,763_ = 25.71, *p* < 0.0001; toxoplasmosis-sex: *F*_2,763_ = 3.84, *p* = 0.022). In the following analyzes, we used the simplest model without Rh and Rh-toxoplasmosis interaction.

The follow-up univariate analysis of covariance (ANCOVA) performed separately for anxiety or sex showed that neither toxoplasmosis nor toxoplasmosis-sex interaction had a significant effect on anxiety (*p* > 0.82). On the other hand, toxoplasmosis-sex interaction had a significant effect on stress (*F*_1,764_ = 6.17, *p* = 0.013, *eta*^2^ = 0.008). Figure 1 shows that the *Toxoplasma*-infected men, but not women had a higher level of stress measured with PSS. Table 1 shows stress, anxiety, physical condition, and mental conditions of particular groups of respondents.

**Table 1.** Perceived stress, anxiety, physical health, and mental health of respondents

|  |  | Stress |  |  | Anxiety |  |  | Physical health |  |  | Mental health |  |  |
| --- | --- | --- | --- | --- | --- | --- | --- | --- | --- | --- | --- | --- | --- |
|  |  | N | Mean | SD | N | Mean | SD | N | Mean | SD | N | Mean | SD |
| Men | Toxoplasma-free | 135 | 30.9 | 3.3 | 138 | 45.2 | 5.3 | 138 | 4.1 | 1.2 | 138 | 4.1 | 1.4 |
| Men | Toxoplasma-infected | 18 | 32.8 | 2.4 | 18 | 44.4 | 4.2 | 18 | 3.8 | 1.2 | 17 | 3.9 | 1.0 |
| Women | Toxoplasma-free | 473 | 32.3 | 3.2 | 476 | 46.4 | 4.9 | 476 | 3.9 | 1.2 | 475 | 3.9 | 1.3 |
| Women | Toxoplasma-infected | 143 | 32.0 | 3.3 | 144 | 45.8 | 4.5 | 144 | 3.8 | 1.2 | 144 | 3.7 | 1.3 |

**Figure 1.**
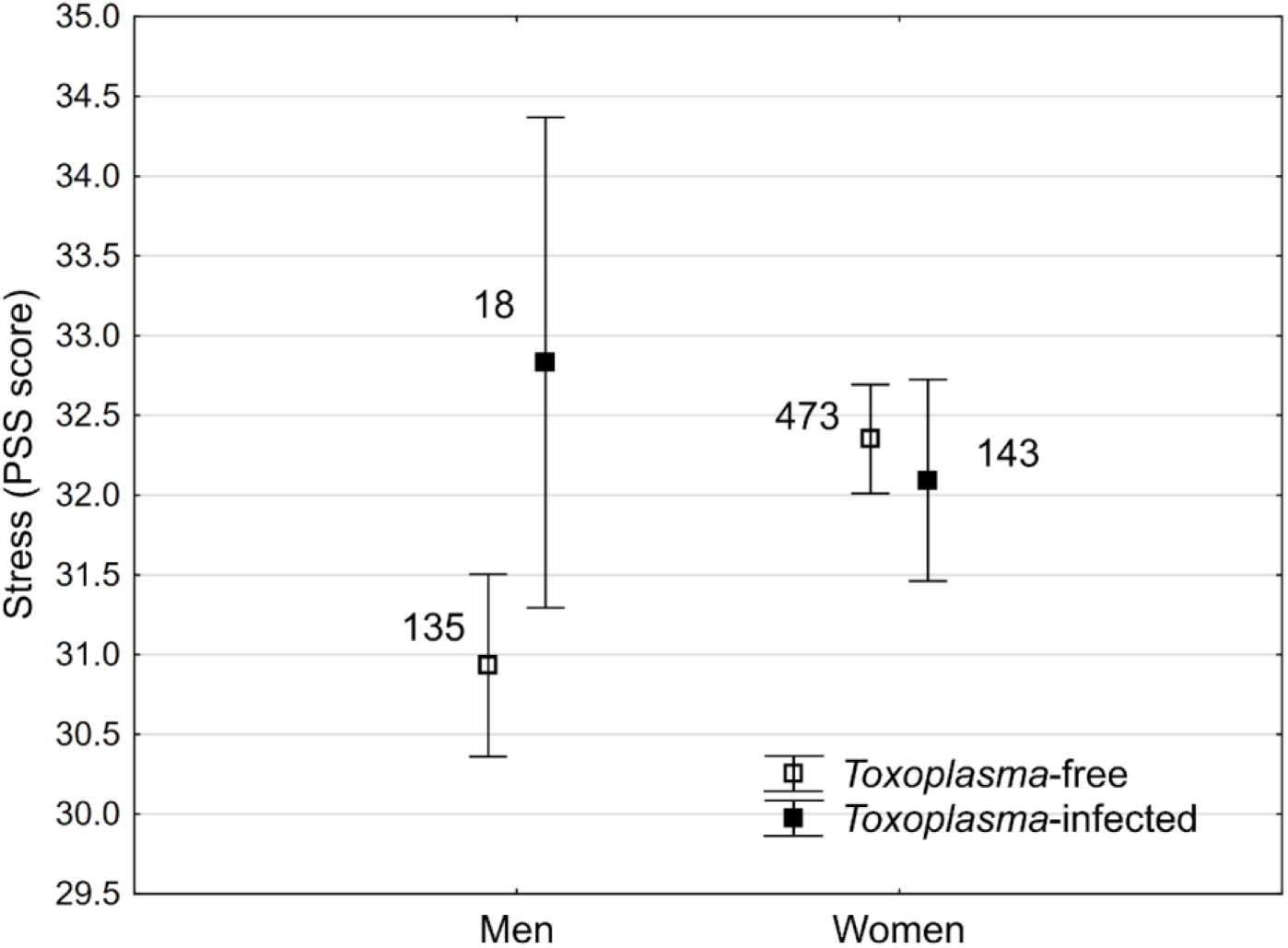
Effect of toxoplasmosis on stress in men and women *Vertical bars denote 0*.*95 confidence intervals, the squares show mean response computed for covariates at their means – i*.*e. for the age 35*.*2*.

It is known that toxoplasmosis has many cumulative adverse effects on the health of infected subjects. Therefore, we also included two other ordinal variables, physical and mental health (higher scores mean better health) as covariates into the analyzed models. The ANCOVA tests showed that higher anxiety was significantly associated with worse mental health (*F*_1,767_ = 110.88, *p* < 0.0001, *eta*^2^ = 0.126, *beta* = -0.384) and lower age (*F*_1,767_ = 24.40, *p* < 0.0001, *eta*^2^ = 0.031, *beta* = -0.165), but not with physical health (*F*_1,767_ = 0.235, *p* = 0.638, *eta*^2^ = 0.0003, *beta* = 0.017) or toxoplasmosis (*F*_1,767_ = 0.345, *p* = 0.557, *eta*^2^ = 0.0005, *beta* = 0.030), or the toxoplasmosis-sex interaction (*F*_1,767_ = 0.027, *p* = 0.870, *eta*^2^ < 0.0003, *beta* = -0.010). In contrast, higher stress covaried with worse mental health (*F*_1,760_ = 90.403, *p* < 0.0001, *eta*^2^ = 0.106, *beta* = -0.362), better physical health (*F*_1,760_ = 5.201, *p* = 0.023, *eta*^2^ = 0.007, *beta* = 0.086) and the toxoplasmosis-sex interaction (*F*_1,760_ = 7.444, *p* = 0.007, *eta*^2^ = 0.010, *beta* = -0.168), but not age (*F*_1,760_ = 2.776, *p* = 0.098, *eta*^2^ = 0.004, *beta* = 0.05) or toxoplasmosis (*F*_1,760_ = 2.747, *p* = 0.094, *eta*^2^ = 0.004, *beta* = -0.087). The correlation of stress with physical health was positive when mental health was controlled in this full model. However, in a simplified model not containing the covariate mental health, this correlation was negative – i.e., the subjects with worse physical health expressed a higher level of stress (*F*_1,761_ = 3.962, *p* = 0.047, *eta*^2^ = 0.005, *beta* = -0.071).

### 3.3 Sex differences in the influence of toxoplasmosis on anxiety and stress

To examine the nature of the toxoplasmosis-sex interaction we analyzed men and women separately with partial Kendall tests. The results showed that toxoplasmosis correlated with stress in men (*Tau* = 0.179, *p* = 0.001) but not in women (*Tau* = -0.028, *p* = 0.304); no association between toxoplasmosis and anxiety was significant. Separate analyses for Rh-positive and Rh-negative men showed that the toxoplasmosis-stress association was of similar strength in Rh-positive men (*Tau* = 0.174, *p* = 0.005) and Rh-negative men (*Tau* = 0.190, *p* = 0.115), however, the association was significant only in more numerous Rh-positive men (118 vs 35). Separate analyses for Rh-positive and Rh-negative women showed no significant association between toxoplasmosis and stress (both *p* values > 0.272). Analogical separate analyses for Rh-positive and Rh-negative subjects found no significant effect of toxoplasmosis on anxiety in men or women (all *p* values > 0.360).

### 3.4 Association of mental and physical health with anxiety and stress

Partial Kendall correlation controlled for age or age and sex showed that worse mental health correlated strongly with anxiety and stress in both men and women (anxiety men: *Tau* = -0.363, stress men: *Tau* = -0.331, anxiety women: *Tau* = -0.271, stress women: *Tau* = -0.231, all *p* < 0.0001). Worse physical health was correlated only with higher anxiety in women (*Tau* = -0.101, *p* = 0.0002). Partial Kendall correlation also showed that anxiety positively correlated with stress (men: *Tau* = 0.375, *p* < 0.0001; women: *Tau* = 0.290, *p* < 0.0001).

### 3.5 Association between latent toxoplasmosis and mental respectively physical health

Toxoplasmosis correlated significantly with mental health in women (*Tau* = -0.065, *p* = 0.015) but not in men (*Tau* = -0.074, *p* = 0.171). Further analyses split by Rh phenotype, however, showed that the negative association between toxoplasmosis and mental health was significant only in Rh-negative subjects (Rh-negative women: *Tau* = -0.131, *p* = 0.025; Rh-positive women: *Tau* = -0.049, *p* = 0.111). The association between toxoplasmosis and mental health was of a similar strength in Rh-negative women and Rh-negative men (women: *Tau* = -0.131; men: *Tau* = -0.136) but achieved a level of statistical significance only in more numerous women (women: *p* = 0.025, *n* = 137; men: *p* = 0.251, *n* = 36). In Rh-positive subjects the strength of the toxoplasmosis-mental health association was weaker in women than in men (women: *Tau* = -0.049; men: *Tau* = -0.064), but was significant neither in women, nor men (women: *p* = 0.111, N = 483; men: p = 0.301, N = 119). The association between toxoplasmosis and physical health was significant neither in women (*Tau* = -0.018, *p* = 0.509) nor men (*Tau* = -0.052, *p* = 0.334). No significant association between toxoplasmosis and physical health was revealed in the analyses split by Rh-phenotype (all *p* values > 0.139).

## 4. Discussion

We found a significant association between perceived stress measured with PSS and toxoplasmosis-sex interaction and no association between anxiety measured with STAI X-2 and toxoplasmosis. *Toxoplasma*-infected men had a significantly higher level of stress while *Toxoplasma*-infected women had a non-significantly lower level of stress than corresponding *Toxoplasma*-free controls. We found no effect of Rh factor phenotype or its interaction with toxoplasmosis on anxiety or stress. The effect of toxoplasmosis on stress was not mediated by impaired physical or mental health conditions as it was even stronger when the effect of physical and mental health was controlled. In accordance with the bulk of previous results (Flegr & Escudero, 2016; Flegr & Horáček, 2020; Flegr, et al., 2014; Šebánková & Flegr, 2017), toxoplasmosis was associated with impaired health, here mental health, only in Rh-negative subjects.

The increased level of stress in male participants is in an accord with the predictions of the stress-coping hypothesis (Lindová, et al., 2010; Lindová, et al., 2006). The authors of this hypothesis theorized that many behavioral changes observed in *Toxoplasma*-infected subjects are in fact results of mild but long-lasting stress – the result of their impaired health. The impaired health of the *Toxoplasma*-infected subjects was observed in several already published studies (Flegr & Escudero, 2016; Flegr & Horáček, 2020; Flegr, et al., 2014; Flegr, et al., 2020; Šebánková & Flegr, 2017) and was confirmed by this study too. The present study has also brought evidence for an increased level of perceived stress in men with latent toxoplasmosis. There are, however, two problems that complicate the straightforward interpretation of the results of the present study in favor of the stress-coping hypothesis:

First, we did not detect increased stress in women. We can just speculate that the PSS questionnaire, which measures the level of perceived stress rather than the level of real physical stress, better reflects the level of stress in men than in women. Such a gender difference could explain why the correlation between anxiety and stress was stronger in men than in women (0.375 vs. 0.290), however, no independent support for this idea probably exists in scientific literature.

Second, the effect of toxoplasmosis on stress measured with PSS was even more visible when the effect of impaired mental and physical health was controlled. The results even suggest that better physical health results in lower stress when the covariate mental health is controlled, suggesting that those subjects who have objective reasons for stress (here those with worse physical health) are less stressed than those without such reasons.

We have no explanation as to why we found an effect of toxoplasmosis on stress but not on anxiety. Anxiety is the most typical emotional response to long, mild, non-predictable stress, including physical stress (Shin & Liberzon, 2010). A fundamental distinction exists between state anxiety, which is a transitory emotional condition, and which is measured with STAI X-1, and trait anxiety, which is a stable personality characteristic regarding the potential for manifesting state anxiety and which is measured with the STAI X-2 (Endler & Parker, 1990). We searched for a long-term attunement of participants in the study and not for a transient state at the time of participation in the study. Therefore, we expected that the personality trait anxiety would better discriminate between *Toxoplasma*-infected and *Toxoplasma*-free subjects. It is possible that this approach was not optimal – the trait anxiety might be determined mostly genetically or might crystalize early in the development of human personality and cannot be affected later by acquired infection.

In contrast to most previous results (Flegr, Novotná, Fialová, Kolbeková, & Gašová, 2010; Flegr, et al., 2008; Flegr, Sebankova, Priplatova, Chvatalova, & Kankova, 2018; Kaňková, Šulc, & Flegr, 2010; Novotná, et al., 2008), we did not find an effect of toxoplasmosis-Rh interaction on anxiety and stress. We only confirmed this effect on mental health, as the association between toxoplasmosis and mental health was detected only in Rh-negative subjects. About 15 published studies have shown that Rh-negative subjects are more prone to the adverse effects of toxoplasmosis as well as to other negative factors, such as aging, smoking, and fatigue (Flegr, Geryk, Volny, Klose, & Cernochova, 2012; Kaňková, et al., 2010). A recent study, however, showed that Rh-positive heterozygotes have superior health and performance while Rh-positive homozygotes have, in many respects, worse performance than Rh-negative subjects (Flegr, et al., 2020). It is possible to study the effects of Rh-phenotype with an electronic questionnaire method, as a large fraction of people know whether they have Rh-positive or Rh-negative blood. In contrast, only a negligible number of people know whether they are Rh-positive heterozygotes or homozygotes. This can be recognized only by molecular genotyping. It would be therefore useful to repeat this study with a genotyped population. However, it will be rather difficult and expensive to genotypize a large enough population of volunteers.

It was suggested in the past that observed behavioral differences between the infected and non-infected subjects are the cause, rather than the effect of the *Toxoplasma* infection (Robertson, 1965). The results of longitudinal studies (Flegr, et al., 2000; Flegr, et al., 1996) as well as the observation of analogical changes in the behavior of artificially infected laboratory rodents (Hodková, Kodym, & Flegr, 2007; Skallová, Kodym, Frynta, & Flegr, 2006; Vyas, Kim, Giacomini, Boothroyd, & Sapolsky, 2007; Vyas, Kim, & Sapolsky, 2007) demonstrated that this explanation cannot be valid. It is not clear, however, whether observed behavioral and personality changes are (1) the products of the manipulative activity of *Toxoplasma* aimed to increase the chances of transmission from intermediate to definitive host, or (2) the side effects of other manipulative activities of *Toxoplasma* aimed to suppress or redirect immune reactions of the infected host, or (3) side-effects of pathological processes running in the infected organism, or (4) the adaptive or (5) maladaptive reactions of the host to the parasitic infection. It is highly probable that all of these alternatives are valid and can explain specific differences in the behavior and personality between infected and non-infected humans, for the detailed discussion of the topic see (Flegr, 2013).

Theoretically, some unknown factors affecting the probability of *Toxoplasma* infection, e.g. inborn or acquired immunodeficiency, could be responsible for the association between stress or anxiety and toxoplasmosis. This possibility cannot be excluded by any observational study, but only by an experiment – artificial infection of randomly selected hosts. Such experiments cannot be done on humans. There are, however, many results from studies performed on laboratory rodents showing that the toxoplasmosis-associated behavioral changes, including stress and anxiety are caused by the infection, not by some other factor.

The main limitation of the present study was that the participants were self-selected and therefore they did not represent a typical Czech population. This is a more or less serious problem of all studies conducted in accord with the Declaration of Helsinki, i.e., in all studies in which the participants are informed that their involvement is voluntary, and they can decline to participate in it at any time. It is always necessary to keep in mind that the conclusions of studies might hold for a specific segment of population, not for the general population as a whole.

The participants also reported their toxoplasmosis status and Rh-phenotype themselves. In the previous internet study (Flegr, 2017), we checked the toxoplasmosis status provided by 3,827 participants who had been examined in our laboratory within the past 10 years. We found 99.5% correspondence of the information provided by the participants and that recorded in our database (Flegr, 2017). However, about 60% of male and 70% of female participants recruited by the Facebook-based snowball method were tested for toxoplasmosis elsewhere, mostly in relation to their health problems (49.4% of men), or to their pregnancy (37.6% of women) (Flegr & Preiss, 2019). It is highly probable that some subjects misreported whether they are *Toxoplasma*-infected (or Rh-negative) or not. Similarly, some respondents who were *Toxoplasma*-negative during their serological test could have acquired the infection in the time between the serological test and participation in the present study. It is, however, important to remember that the presence of misdiagnosed subjects in the population can result in a Type 2, not a Type 1 error – it can increase the risk of failure to detect existing effects but not the risk of detecting non-existing effects. Theoretically, the stochastic errors could explain the negative result of the test searching for the effect of toxoplasmosis on anxiety. However, we consider this possibility rather improbable as we find a negative, rather than positive, relation between toxoplasmosis and anxiety.

The size of the observed effects of toxoplasmosis on stress might seem relatively low. However, the Kendall Tau 0.190 corresponds to Cohen d 0.31, which is usually classified halfway between small and medium effects.

## 5. Conclusions

In the present study, we showed that *Toxoplasma*-infected men had a higher level of perceived stress than *Toxoplasma*-free men. At face value, this observation provides new support for the stress-coping hypothesis explaining the reason why the toxoplasmosis-associated personality and behavioral shifts go mostly in opposite directions in men and women. However, the absence of this effect in women, as well as the negative effect of physical health problems on the level of stress (when mental health is controlled) indicates that we must be careful with the interpretation of new results. Possibly, we should also rethink whether perceived stress correctly reflects the real level of physical stress. Generally, our results suggest that direct measurement of the concentration of stress hormones might be a better approach for testing the stress-coping hypothesis than measuring perceived stress using psychological instruments.

The effect of latent toxoplasmosis on the health of people was not the topic of the present study. Still, the increased level of perceived stress in men, as well as worse mental health in *Toxoplasma*-infected Rh-negative men and women, brought the new evidence that toxoplasmosis (affecting one-third of the human population) could be a serious and underestimated public health problem.

## Data Availability

All data are available at Fishare repository, doi.org/10.6084/m9.figshare.14651490.v1 (Flegr, 2021)

## Acknowledgments

We would like to thank Lincoln Cline for her help with preparing the final version of the article.

## Statement of Ethics

The project, including the method of obtaining informed consent to participate in this anonymous study from all participants by pressing the corresponding button on the first page of the questionnaire, was approved by the IRB of the Faculty of Science, Charles University (Komise pro práci s lidmi a lidským materiálem Přírodovědecké Fakulty Univerzity Karlovy) — No. 2019/30.

## Author Contributions

Conceptualization, original draft preparation, funding acquisition, and supervision, J.F.; formal analysis, writing—review and editing, investigation J.F. and Š.K. Both authors have read and agreed to the published version of the manuscript.

## Data Availability Statement

All data are available at Fishare repository, doi.org/10.6084/m9.figshare.14651490.v1 (Flegr, 2021)

